# Change in Acetylcholinesterase Activity from Childhood to Young Adulthood

**DOI:** 10.1101/2024.10.21.24315881

**Authors:** Jose R. Suarez-Lopez, Carlos F. Gould, Devesh Vashishtha, Asa Bradman, Jose Suarez-Torres, Dolores Lopez-Paredes, Danilo Martinez, Raeanne M. Moore

## Abstract

**Objective:** Acetylcholinesterase (AChE) is an enzyme that metabolizes acetylcholine, an essential neurotransmitter, and is frequently used to monitor adult agricultural workers for exposure to cholinesterase inhibitor pesticides. Yet, there are no clear standards for AChE activity in children and adolescents, which prohibits evaluations of dangerous pesticide exposures in younger populations.

**Methods:** We measured AChE activity from a single finger stick blood sample data from 746 participants ages 4 to 26 years across 3,100 observations who resided in an agricultural county in Ecuador. We used generalized estimating equations to predict AChE activity levels in one year age increments from 5 to 25 years, accounting for nonlinear aging patterns and survey wave specific effects. We also decomposed variation in observed AChE activity levels into aging effects, differences in our recruited participants, and participant-specific aging patterns.

**Results:** Average AChE activity levels across all observations were 3.88 U/mL (standard deviation [sd] = 0.67). AChE activity levels increased nonlinearly as participants aged. We found that males had higher AChE activity levels than females and that those levels established themselves later in age than females. AChE activity levels increased essentially linearly from ages 5 to 12 or 18 years, depending on sex, at which point levels did not meaningfully change with age. Most variation observed in AChE activity levels were due to aging effects.

**Conclusion:** Our findings provide reference levels for AChE activity across childhood, adolescence, and into early adulthood that can be used by clinicians and researchers in the context of assessing neurodevelopment and potential exposure to neurotoxicants.

---

Acetylcholinesterase (AChE) plays a crucial role in the cholinergic system by hydrolyzing acetylcholine, thus regulating nerve impulse transmission in the peripheral and central nervous systems^1^. Inhibition of AChE activity by commonly used insecticides—such as organophosphates (OPs) and carbamates—as well as certain nerve agents, leads to the accumulation of acetylcholine, resulting in significant physiological alterations and neurotoxicity^23^. OPs are among the most common causes of poisoning worldwide, stemming from agricultural, accidental, or suicidal exposures. Due to the prevalence and severity of OP toxicity, monitoring potentially toxic exposures is essential in agricultural settings—frequently achieved by comparing pre-exposure (basal) AChE activity levels to levels during or shortly after pesticide application. For instance, in states like Washington and California, a formal workplace evaluation is required if AChE levels drop below 80% of basal activity^10^. In situations where regular basal monitoring is not conducted, reference values are crucial for comparing an individual’s AChE activity to expected levels in the general population. Since AChE activity levels serve as indicators of both neurological function and pesticide exposure, quantifying AChE levels across different ages and populations is vital for understanding normal physiological development and identifying potential disruptions caused by environmental exposures such as pesticides.

While adults typically have stable AChE activity over time^4–6^, the trajectory of AChE activity levels from childhood to adulthood remains poorly quantified. Karlsen et al. (1981)^7^ studied 217 children aged 0 to 12 years and 30 adults in Norway, finding that AChE activity increases rapidly during the first few months of life and reaches approximate adult levels by one year of age (around 10 units per gram of hemoglobin [U/g Hb]). However, their estimates for each age were based on small sample sizes (8–10 participants per age), limiting the precision of their findings. In a cross-sectional sample of 277 participants recruited from rural agricultural Ecuador, we previously found a linear increase in AChE activity by 0.05 U/mL per year from ages 4-9 years^8^. Worek et al. (2016)^9^ examined AChE levels in 242 adults aged 18 to 61 years in Germany, observing minimal changes in AChE activity across ages in young to middle-aged adults; however, this study was also cross-sectional and did not follow participants over time. The lack of longitudinal data on AChE activity from childhood through young adulthood represents a significant gap in the literature.

Establishing age-specific reference values for AChE is essential for clinicians to distinguish normal physiological development from potential neurotoxic effects resulting from environmental exposures such as pesticides. These benchmarks enable early detection and intervention, mitigating long-term health impacts and informing public health policies to protect vulnerable populations, particularly children in agricultural communities. This study aims to address these knowledge gaps by characterizing AChE activity across key developmental stages from childhood through early adulthood, providing much-needed age-specific AChE activity standards.

## Materials and Methods

### Participants and Study design

The study of Secondary Exposures to Pesticides among Children, Adolescents and Adults (ESPINA) is a prospective cohort study established in 2008 to investigate the effects of pesticide exposures on human development. ESPINA sought to have a balanced distribution of participants living with a flower plantation worker and participants not living with any agricultural workers. To be eligible to participate, participants must have either (a) lived with a flower plantation worker for at least one year or (b) never lived with an agricultural worker, never inhabited a house where agricultural pesticides were stored, and have had no previous direct contact with pesticides.

The cohort has been described previously^8^. We examined 313 boys and girls aged 4-9 years who lived in Pedro Moncayo, Pichincha province, Ecuador in 2008. New participants were recruited from a local community census in 2016 (n=314) and 2022 (n=121). In 2023 and 2024, four additional examinations were conducted, during which no new participants recruited. Participants were examined in schools in all five parishes of Pedro Moncayo County.

### Data Collection

#### Anthropometric measures

Height was measured to the nearest 1 mm using a stadiometer following recommended procedures ^23^, and weight was measured using digital scales.

#### Acetylcholinesterase and hemoglobin

Erythrocytic AChE activity and hemoglobin concentrations (mg/dL) waas measured with the EQM Test-mate ChE Cholinesterase Test System 400 (EQM AChE Erythrocyte Cholinesterase Assay Kit 470; EQM Research, Inc, Cincinnati, OH) from a single finger-stick blood sample, following standard procedures ^24^.

#### Urinary pesticide metabolites

Urinary concentrations of organophosphate metabolites, including malathion dicarboxylic acid (MDA), 3,5,6-Trichloro-2-pyridinol (TCPy), and para-Nitrophenol (PNP), were quantified. The target metabolites were extracted from 1.5 mL of urine using a semi-automated solid-phase extraction method. Separation from other urinary components was achieved using reversed-phase high-performance liquid chromatography, and the metabolites were identified using tandem mass spectrometry with isotope dilution quantitation. Strict quality control procedures were followed throughout to ensure accuracy, precision, reproducibility, and efficiency of the method. Metabolites were measured at the Centers for Disease Control and Prevention, Division of Laboratory Sciences. The limits of detection for the metabolites were 0.5 µg/L for MDA and 0.1 µg/L for TCPy and PNP.

#### Urine specific gravity

Specific gravity in 2022 and 2024 was measured in fresh urine samples using an ATAGO PAL-10S Urine Specific Gravity Refractometer (Atago Co., Tokyo, Japan), or an ATAGO Wrestling Digital Hand-held PEN Urine Specific Gravity Refractometer (Atago Co., Tokyo, Japan). Samples collected in January-February 2023 and July 2023 were measured using previously frozen urine samples (stored at −20°C), which were thawed and brought to room temperature before analysis.

### Statistical analysis

We used generalized estimating equations (GEE) to model the relationship between AChE levels and various predictors, accounting for repeated measures. Predictors included age (linear and quadratic terms for non-linear effects), sex, hemoglobin, height, BMI, a fourth-degree polynomial for exam date (to capture seasonal pesticide exposures), and exam wave. We specified an exchangeable correlation structure to address within-subject correlations. To generate smooth, age-specific reference values, we employed a two-stage prediction process. First, we modeled hemoglobin, height, BMI, and exam date as functions of age and sex using quartic splines in generalized additive models (GAMs), capturing non-linear age trends by sex. Then, using the GEE model and smoothed covariate estimates, we predicted AChE activity for ages 4 to 26 in 0.1-year increments for both sexes and all exam waves. We averaged predictions across exam waves for each age-sex group and then across sexes to obtain population-averaged reference values. This approach assumes similar AChE progression across sexes. To relax this, we interacted sex with linear and quadratic age terms and repeated the process to generate sex-stratified predictions.

To address the concern that our findings are influenced by the composition of our sample, we conduct a bootstrapping procedure whereby we sample individuals with replacement and re-analyze our data. This approach simulates random sampling from a larger super population 1,000 times. We report central estimates as the mean of these estimates and the 2.5^th^ to the 97.5^th^ percentile of estimates.

To identify the age at which trends in AChE activity levels diverged across females and male, we used the ‘slopes’ function from the ‘marginaleffects’ package to generate predictions and identify sex-specific differences at each age interval. Then we identified the age at which predicted sex-specific AChE levels were statistically significantly different (at P<0.05).

We aimed to determine how much of the changes in AChE levels across ages were due to within-individual aging versus persistent between-individual differences in our sample. To do this, we fit a mixed-effects model with fixed and random effects for age, including a random intercept for each individual (accounting for between-individual differences) and a random slope for age (capturing individual-specific aging trajectories). By comparing the variances of these random effects, we assessed the relative contributions of within- and between-individual differences. We extracted three key variance components: (a) residual variance (within-individual changes), (b) random intercept variance (between-individual differences in overall levels), and (c) random slope variance (between-individual differences in aging rates). The proportion of variance attributable to each component helped us determine how much of the variation in AChE levels was due to within-individual aging, between-individual differences in overall levels, and between-individual differences in aging rates.

Previous evidence has established that AChE activity levels are associated with urinary organophosphate pesticide metabolites ^11^; furthermore, our study population is in an agricultural setting where participants may have exposures to such pesticides. Unfortunately, we did not collect pesticide metabolites in every study wave, so we are unable to directly control for pesticide exposures in our main model. Given these constraints, we aimed to identify the influence of pesticide exposures in our *predictions* of AChE activity levels across ages. To do so, we predicted AChE activity levels across ages while directly including PNP, TCPy, and MDA concentrations (available for waves C and D) and their squares as additional controls in regression models. We compared these predictions with predictions made based on data from waves C and D without the inclusion of PNP, TCPy, and MDA.

We used R software version 4.4.0 to conduct all analyses; packages used included ‘geepack for GEEs, ‘segmented’ for breakpoint regressions, and ‘mgcv’ for GAMs. Statistical significance was determined at P<0.05.

### Ethics

We acquired informed consent for participation from adults and obtained parental permission for participation of minor children. We also obtained child assent from participants who were between 7 and 18 years of age. The institutional review boards at the University of California San Diego (USA), University of Minnesota (USA), Universidad San Francisco de Quito (Ecuador), UTE University (Ecuador) and the Ministry of Public Health of Ecuador approved this study. Additionally, the Commonwealth of Rural Parishes of Pedro Moncayo County endorsed this project and access to schools was approved by the Ministry of Education.

## Results

### Participant characteristics

**Table 1** presents descriptive statistics for the 747 participants (3122 observations) that had at least one valid AChE activity measure. An average of 390 participants were surveyed in each of the eight exam periods (minimum = 311, maximum = 534). Four-fifths of participants were surveyed at least once, two-thirds at least three times, half at least five times, and one-tenth were surveyed in all eight waves. See **Figure S1** for the distribution of when participants were surveyed by wave.

**Table 1.** Participant characteristics.

|  | **Total**<br>**(N=3122)** | **Wave A**<br>**(N=311)** | **Wave B**<br>**(N=322)** | **Wave C**<br>**(N=533)** | **Wave D**<br>**(N=501)** | **Wave F**<br>**(N=355)** | **Wave H**<br>**(N=374)** | **Wave J**<br>**(N=371)** | **Wave L**<br>**(N=352)** |
| --- | --- | --- | --- | --- | --- | --- | --- | --- | --- |
| **Exam dates** | July 2008-July 2024 | July 2008-August 2008 | April 2016-May 2016 | July 2016-October 2016 | July 2022-August 2022 | January 2023-February 2023 | July 2023 | January 2024-February 2024 | June 2024-July 2024 |
| **Age, years** |  |  |  |  |  |  |  |  |  |
| Mean (SD) | 17.8 (5.1) | 6.6 (1.6) | 14.2 (1.8) | 14.5 (1.8) | 20.3 (1.8) | 20.7 (1.8) | 21.3 (1.8) | 21.7 (1.8) | 22.1 (1.8) |
| Median | 19.4 | 6.5 | 14.1 | 14.4 | 20.1 | 20.6 | 21 | 21.4 | 21.9 |
| Q1,Q3 | 14.6, 21.6 | 5.3, 7.8 | 12.8, 15.7 | 13.1, 15.8 | 18.8, 21.7 | 19.2, 22.1 | 19.7, 22.6 | 20.1, 23.0 | 20.5, 23.5 |
| **Sex** |  |  |  |  |  |  |  |  |  |
| Female | 1671 (53.5%) | 153 (49.2%) | 159 (49.4%) | 270 (50.7%) | 252 (50.3%) | 201 (56.6%) | 213 (57.0%) | 216 (58.2%) | 207 (58.8%) |
| Male | 1451 (46.5%) | 158 (50.8%) | 163 (50.6%) | 263 (49.3%) | 249 (49.7%) | 154 (43.4%) | 161 (43.0%) | 155 (41.8%) | 145 (41.2%) |
| **AChE, U/mL** |  |  |  |  |  |  |  |  |  |
| Mean (SD) | 3.9 (0.7) | 3.1 (0.5) | 3.8 (0.5) | 3.7 (0.5) | 4.4 (0.6) | 4.0 (0.6) | 4.1 (0.6) | 3.9 (0.6) | 4.1 (0.6) |
| Median | 3.9 | 3.1 | 3.8 | 3.7 | 4.4 | 4 | 4.1 | 3.9 | 4 |
| Q1,Q3 | 3.5, 4.3 | 2.8, 3.5 | 3.5, 4.1 | 3.3, 4.1 | 4.0, 4.9 | 3.7, 4.4 | 3.7, 4.4 | 3.5, 4.3 | 3.7, 4.4 |
| **Hemoglobin,** |  |  |  |  |  |  |  |  |  |
| Mean (SD) | 13.7 (1.6) | 12.6 (1.2) | 13.0 (1.3) | 13.0 (1.2) | 14.7 (1.8) | 13.9 (1.6) | 14.1 (1.5) | 13.8 (1.5) | 14.3 (1.5) |
| Median | 13.6 | 12.5 | 13.0 | 12.9 | 14.6 | 13.8 | 14.2 | 13.8 | 14.2 |
| Q1,Q3 | 12.6, 14.7 | 11.9, 13.3 | 12.3, 13.7 | 12.2, 13.6 | 13.5, 15.8 | 13.0, 14.9 | 13.2, 15.1 | 12.8, 14.8 | 13.4, 15.1 |
| **Height, cm** |  |  |  |  |  |  |  |  |  |
| Mean (SD) | 151.8 (16.5) | 112.1 (10.3) | 149.0 (9.6) | 150.6 (9.4) | 159.2 (8.7) | 158.8 (8.8) | 158.8 (8.7) | 159.0 (8.7) | 159.0 (8.6) |
| Median | 154.1 | 112.5 | 149 | 150.5 | 158.7 | 157.5 | 157.7 | 157.7 | 157.8 |
| Q1,Q3 | 147.0, 162.5 | 104.3, 119.8 | 142.4, 154.9 | 145.0, 156.6 | 152.6, 165.4 | 152.0, 165.3 | 152.0, 165.4 | 152.0, 165.1 | 152.8, 165.0 |
| **BMI, kg/m<sup>2</sup>** |  |  |  |  |  |  |  |  |  |
| Missing | 1 | 1 | 0 | 0 | 0 | 0 | 0 | 0 | 0 |
| Mean (SD) | 22.5 (4.1) | 16.1 (1.3) | 20.8 (3.0) | 20.9 (2.9) | 24.0 (3.5) | 24.2 (3.5) | 24.3 (3.5) | 24.4 (3.6) | 24.5 (3.5) |
| Median | 22.4 | 16 | 20.5 | 20.6 | 23.6 | 24 | 23.9 | 23.9 | 24.1 |
| Q1,Q3 | 19.7, 24.9 | 15.3, 16.8 | 18.7, 22.6 | 18.8, 22.4 | 21.5, 25.7 | 21.7, 26.4 | 21.9, 26.1 | 21.8, 26.4 | 22.2, 26.4 |

Roughly half of participants were female in the first four exam waves; in the last four exam waves around 57% of participants were female. In the first exam wave – July 2008-August 2008 – the average age of participants was 6.6 years. During the final exam wave – June to July 2024 – the average age of participants was 22.1 years. Across all waves, the average and median ages of participants at the time of exams was 17.8 and 19.4 years, respectively; a wide range of ages was covered across exam waves: the 1^st^ to 99^th^ percentile of ages was 4.4 to 25.3 years. Owing to the distribution of participants’ sex skewing towards females in later exam waves, females were about 0.7 years older than males on average across the full study sample (**Figure S2**). Still, female and male participants were of similar ages when comparing participants’ ages within waves. The average AChE activity levels across all observations was 3.90 U/mL (SD = 0.67); the 1^st^ to 99^th^ percentiles were 2.42 to 5.48 U/mL. Height, weight, BMI, and hemoglobin concentrations increased across waves as participants aged.

### Age and AChE activity

AChE activity increased with age from childhood through young adulthood (**Figure 1A**), reaching adult levels at 18-20 years of age. The raw median and interquartile range AChE activity levels when participants were 5-10 years of age were 3.19 U/mL (IQR, 2.89 - 3.51; N=253 observations); these values increased to 4.08 U/mL (IQR, 3.68-4.50; N=900 observations) when participants were between 15-20 years of age where they remained unchanged among older ages.

**Figure 1.**
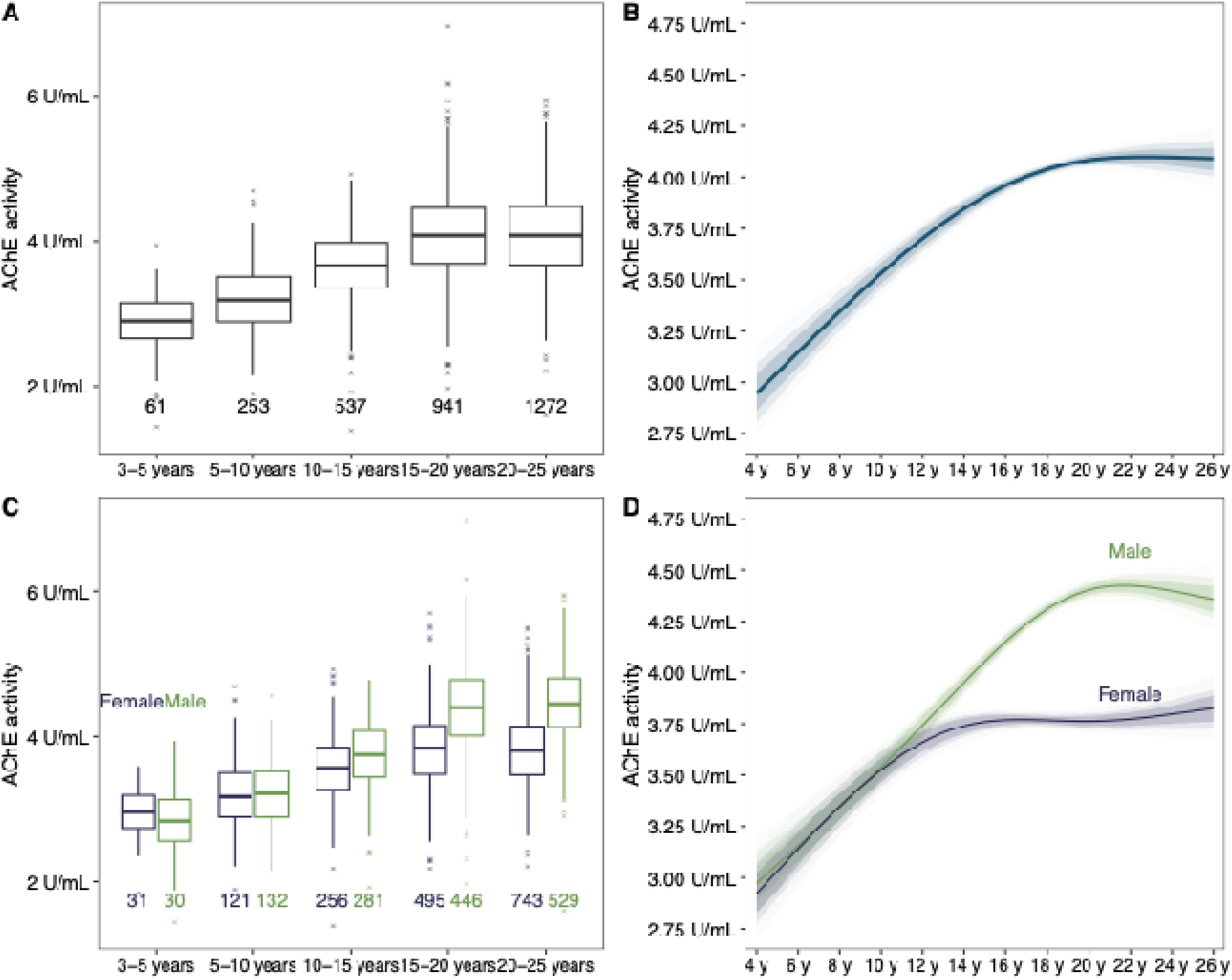
Observed and predicted ACHE activity levels by age and sex. **A** presents AChE activity levels by age group where boxplots show the median, interquartile range, and 1.5 times the interquartile range (IQR) as hinges. Outliers that extend beyond the IQR are annotated with partially transparent Xs. Sample sizes within each group range are shown below the box plots. **B** shows predicted AChE activity levels continuously between ages 4 to 26 years from GEE models where the main prediction is the average of 1000 bootstrapped samples and shaded areas indicate the 80^th^, 90^th^, and 95^th^ percentiles of the distribution at each 0.1-year increment. **C** and **D** indicate the same but stratified by sex. The models presented in **D** directly interact sex with age.

**Figure 1B** and **Table 2** present results from this model as predicted AChE activity levels from 5 to 25 years. AChE activity levels increased linearly between 5 and 15 years, began to flatten between 15 and 20 years, and were steady thereafter through 25 years. At age 5 years, average predicted AChE activity levels across all bootstrapped model runs were 2.92 U/mL, with the 2.5^th^ to 97.5^th^ percentiles being 2.67 U/mL to 3.14 U/mL. Predicted AChE activity levels increased through age 18 years, where average levels were 4.04 U/mL (2.5^th^ to 97.5^th^, 4.00-4.08 U/mL) and then held roughly constant through age 25 years (average 4.10 U/mL, 2.5^th^ to 97.5^th^, 3.98-4.22 U/mL).

**Table 2.** Observed and model predicted AChE activity levels by age.

| Age (y) | Full sample |  |  |  | Females |  |  |  | Males |  |  |  |
| --- | --- | --- | --- | --- | --- | --- | --- | --- | --- | --- | --- | --- |
|  | Observed |  | Predicted |  | Observed |  | Predicted |  | Observed |  | Predicted |  |
|  | N | Mean (U/mL) | Mean (U/mL) | 5<sup>th</sup>-95<sup>th</sup> (U/mL) | N | Mean (U/mL) | Mean (U/mL) | 5<sup>th</sup>-95<sup>th</sup> (U/mL) | N | Mean (U/mL) | Mean (U/mL) | 5<sup>th</sup>-95<sup>th</sup> (U/mL) |
| 5 | 56 | 3.03 | 3.05 | 2.86-3.26 | 32 | 2.93 | 3.03 | 2.83-3.24 | 24 | 3.16 | 3.06 | 2.88-3.28 |
| 6 | 58 | 3.12 | 3.15 | 2.98-3.34 | 28 | 3.13 | 3.14 | 2.95-3.35 | 30 | 3.11 | 3.16 | 2.99-3.35 |
| 7 | 59 | 3.22 | 3.25 | 3.10-3.42 | 26 | 3.31 | 3.25 | 3.09-3.43 | 33 | 3.14 | 3.25 | 3.1-3.42 |
| 8 | 56 | 3.2 | 3.34 | 3.21-3.50 | 26 | 3.19 | 3.35 | 3.2-3.5 | 30 | 3.21 | 3.34 | 3.21-3.49 |
| 9 | 37 | 3.43 | 3.44 | 3.32-3.57 | 17 | 3.43 | 3.44 | 3.3-3.58 | 20 | 3.43 | 3.44 | 3.31-3.58 |
| 10 | 13 | 3.32 | 3.53 | 3.42-3.65 | 5 | 3.03 | 3.53 | 3.41-3.65 | 8 | 3.5 | 3.54 | 3.42-3.66 |
| 11 | 21 | 3.51 | 3.62 | 3.52-3.72 | 9 | 3.49 | 3.6 | 3.49-3.71 | 12 | 3.54 | 3.64 | 3.53-3.75 |
| 12 | 114 | 3.59 | 3.7 | 3.60-3.79 | 57 | 3.66 | 3.66 | 3.56-3.75 | 57 | 3.53 | 3.74 | 3.65-3.84 |
| 13 | 166 | 3.64 | 3.78 | 3.69-3.85 | 87 | 3.55 | 3.71 | 3.61-3.79 | 79 | 3.74 | 3.85 | 3.77-3.93 |
| 14 | 155 | 3.72 | 3.85 | 3.77-3.91 | 68 | 3.47 | 3.74 | 3.65-3.81 | 87 | 3.91 | 3.95 | 3.87-4.02 |
| 15 | 153 | 3.81 | 3.91 | 3.84-3.96 | 77 | 3.74 | 3.76 | 3.69-3.82 | 76 | 3.88 | 4.05 | 3.99-4.11 |
| 16 | 123 | 3.94 | 3.96 | 3.90-4.01 | 65 | 3.74 | 3.77 | 3.71-3.82 | 58 | 4.16 | 4.15 | 4.09-4.2 |
| 17 | 96 | 3.95 | 4.00 | 3.96-4.04 | 50 | 3.62 | 3.77 | 3.72-3.82 | 46 | 4.32 | 4.24 | 4.18-4.29 |
| 18 | 161 | 4.24 | 4.04 | 4.00-4.08 | 78 | 3.90 | 3.77 | 3.73-3.81 | 83 | 4.57 | 4.31 | 4.26-4.37 |
| 19 | 290 | 4.19 | 4.06 | 4.02-4.10 | 146 | 3.92 | 3.76 | 3.72-3.81 | 144 | 4.45 | 4.36 | 4.31-4.42 |
| 20 | 378 | 4.12 | 4.08 | 4.04-4.13 | 209 | 3.85 | 3.76 | 3.71-3.82 | 169 | 4.46 | 4.4 | 4.34-4.46 |
| 21 | 334 | 4.02 | 4.09 | 4.04-4.15 | 200 | 3.76 | 3.77 | 3.7-3.84 | 134 | 4.41 | 4.42 | 4.35-4.48 |
| 22 | 298 | 4.1 | 4.10 | 4.03-4.17 | 170 | 3.82 | 3.77 | 3.69-3.86 | 128 | 4.47 | 4.42 | 4.34-4.49 |
| 23 | 246 | 4.1 | 4.10 | 4.02-4.19 | 149 | 3.87 | 3.78 | 3.69-3.89 | 97 | 4.46 | 4.41 | 4.32-4.51 |
| 24 | 162 | 4.08 | 4.10 | 4.00-4.20 | 98 | 3.83 | 3.8 | 3.68-3.92 | 64 | 4.46 | 4.4 | 4.29-4.51 |
| 25 | 85 | 4.01 | 4.09 | 3.98-4.22 | 47 | 3.61 | 3.81 | 3.68-3.95 | 38 | 4.51 | 4.38 | 4.25-4.51 |

### Age and AChE activity stratified by sex

**Figure 1C** presents AChE levels by sex and age group. Across the full sample, when controlling for age, hemoglobin, height, BMI, and exam wave, AChE activity levels were 0.13 U/mL higher among males (95% confidence interval [CI]: 0.06-0.19 U/mL; 3.2% higher [95% CI: 1.5%-5.0%]). **Figure 1D** and **Table 2** present results from a model as predicted AChE activity levels from 5 to 25 years for males and females separately. Predicted average AChE activity levels were similar for both males and females from ages 5 to 12 years. At age 5 years, predicted average AChE activity levels were 2.91 U/mL (2.5^th^ to 97.5^th^, 2.68-3.12 U/mL) and 2.95 U/mL (2.5^th^ to 97.5^th^, 2.69-3.19 U/mL) for females and males, respectively. From ages 12 to 25 years, however, we observed significant differences in AChE activity levels according to sex. Differences between predicted AChE acitivity between male and female participants appear between 10 and 11 years of age; at age 12.5 years, analytic confidence intervals are statistically different between the sexes (**Figure S3**). Still, AChE activity levels are increasing at this age for both male and female participants. Predicted AChE activity levels among males continued to increase through age 19 years when they leveled off around 4.40 U/mL. Among females, however, AChE activity levels leveled off closer to age 15 years at 3.75 U/mL.

### Decomposition of variation

We find that 64% of the variation observed in AChE activity levels are attributable to within-subject differences, which we interpret as ‘the effect of aging.’ The remaining variation is largely attributable to between-subject differences, i.e., ‘differences across individuals.’ Less than 1% of our observed variation is due to differences in aging rates across individuals.

### Robustness to inclusion of pesticide metabolites

We find no meaningful influence of the inclusion of pesticide metabolites – PNP, TCPy, and MDA – on predicted AChE activity levels between ages 12 and 23 years (the 5^th^-95^th^ percentiles of ages in waves C and D) (**Figure S5**). When we set each of these pesticide metabolite levels to their 10^th^ percentile, predicted AChE activity levels were marginally higher than when pesticide metabolite levels were set to their 90^th^ percentiles – as we would expect – though point estimates were still very similar (**Figure S6**).

## Discussion

We present overall and sex-specific AChE activity levels from ages 5 to 25 years, using over 3,100 samples from more than 700 participants collected across eight exam waves between 2008 and 2024. Males and females had similar AChE activity levels between ages 4 to 10 years during which point they linearly increased from about 3.0 U/mL to 3.5 U/mL. After 10 years of age, sex-specific patterns emerged. In males, AChE activity continued to rise quasi-linearly until age 18, leveling off around 4.40 U/mL. In females, AChE activity plateaued earlier, around age 12, at a lower level of approximately 3.75 U/mL. These age-related changes in AChE activity are largely attributable to aging, rather than variations in the participant sample across exam waves.

These findings align with existing knowledge on the timing of puberty, where females mature earlier than males, and with animal studies showing similar early-age AChE levels but higher levels in older males. While animal studies suggest aging as a key driver of AChE changes, previous human studies relied on cross-sectional samples. Our longitudinal data, however, allow us to quantify within-individual changes over time, confirming that the observed AChE trends are primarily driven by aging rather than sample composition effects.

Given the established harmful effects of organophosphate insecticides, the WHO has recommended that agricultural workers who handle cholinesterase inhibiting pesticides or who work in fields sprayed with these chemicals should be monitored for blood cholinesterase levels between pre- and post-exposure measurements ^25^. Field studies throughout the world have shown that biomonitoring in adults helps detect and prevent pesticide poisoning ^26–29^. In these cases, clinicians and toxicologists may draw on reference values for AChE activity levels typical among adults to samples collected from potentially exposed individuals. Given that many young adults, and even adolescents and children, may either directly participate in agricultural work, live near spraying sites, or live with individuals that are involved in spraying, these reference values are needed for this vulnerable population. We provide such values and indicate that, at least in our sample, females aged 13 years and older and males aged 19 years and older may have AChE activity levels similar to those found in adults.

Although red blood cell (RBC) AChE activity is less sensitive and specific than urinary pesticide metabolite quantification, it offers practical advantages for screening pesticide exposure. It is 10–15 times cheaper, requires only a drop of blood, and provides results within 5– 6 minutes. RBC AChE may also reflect exposure over a longer time window and may be more stable than urinary metabolites, which can fluctuate due to hydration and recent exposures. A criticism of using RBC AChE is its low index of individuality, around 0.45, suggesting limited utility of population-based reference intervals. However, many common clinical analytes like insulin, creatinine, cortisol, and cholesterol have similar index of individuality values. This suggests population reference intervals for AChE remain useful. Our findings illustrate the applicability of RBC AChE activity as a one-time screening tool for pesticide exposure in children, especially in settings where more sensitive assays are not feasible.

Our work is subject to some limitations. Our participants had a higher potential for exposure to pesticides than the general population of Ecuador, given that much of the economy of Pedro Moncayo County is based on agriculture. We sought to minimize the influence of potential direct or indirect exposures to cholinesterase inhibitors by sampling during periods of low flower production and pesticide use – the floricultural industry reduces its production of flowers during the summer and early fall due to an absence of holidays with big flower sales during that time. While this was a non-trivial challenge because our study setting that permits year-round agricultural production, we were able to do so thanks to high community interest and involvement. Still, it is plausible that the AChE levels among study participants may have been lower than those of a similar set of participants that lived further from agriculture. Nevertheless, predicted AChE activity levels were nearly identical when modeled at low and high pesticide metabolite levels among the subset of participants with measures. Future studies should conduct similar analyses within populations with a lower potential for pesticide exposures. Nevertheless, our sample – children and young adults living in agricultural communities – are a major target pesticide exposure screening using AChE activity, and thus our reference values may be particularly applicable for others conducting exams in similar settings.

## Conclusion

We provide reference values for age-based standards of AChE levels in children, adolescents, and young adults, which can be used for epidemiological studies, clinical evaluation, and pesticide exposure screening. Further longitudinal studies might involve similar analyses among populations of children, adolescents, and young adults with a lower risk of pesticide exposure.

## Data Availability

Private use data in this study are not publicly available.

## Acknowledgments

We are indebted to the ESPINA study participants, without whom this work would be impossible.

## Abbreviations

AChE: acetylcholinesterase
OP: organophosphate
ESPINA: Secondary Pesticide Exposure among Children and Adolescent
GEE: generalized estimating equations
BMI: body mass index
GAM: generalized additive model
TCPy: 3,5,6-trichloro-2-pyridinol
PNP: 4-Nitrophenol
MDA: malathion dicarboxylic acid
SD: standard deviation
CI: confidence interval

## SUPPLEMENTAL INFORMATION

**Figure S1.**
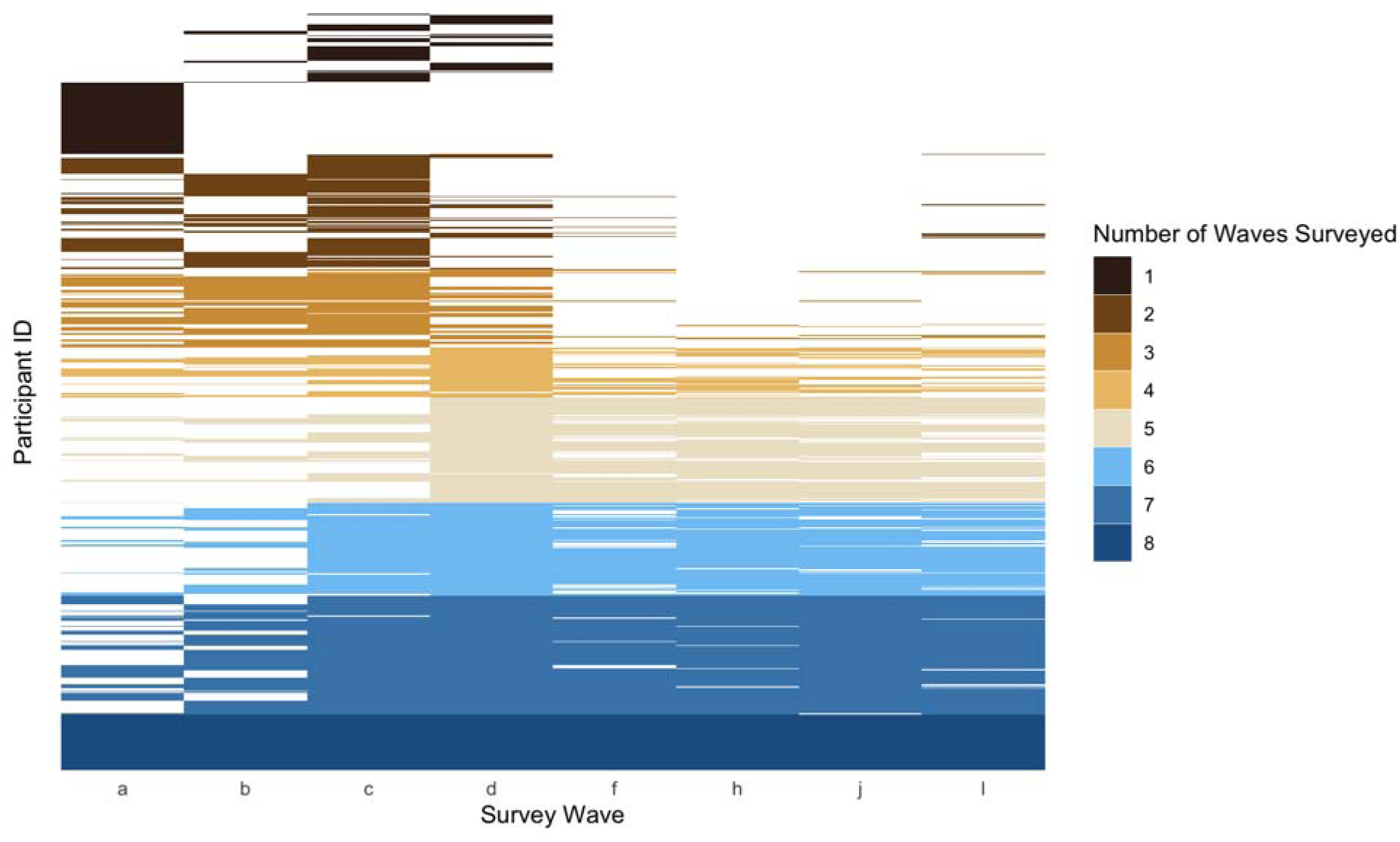
Distribution of waves when participants were surveyed.

**Figure S2.**
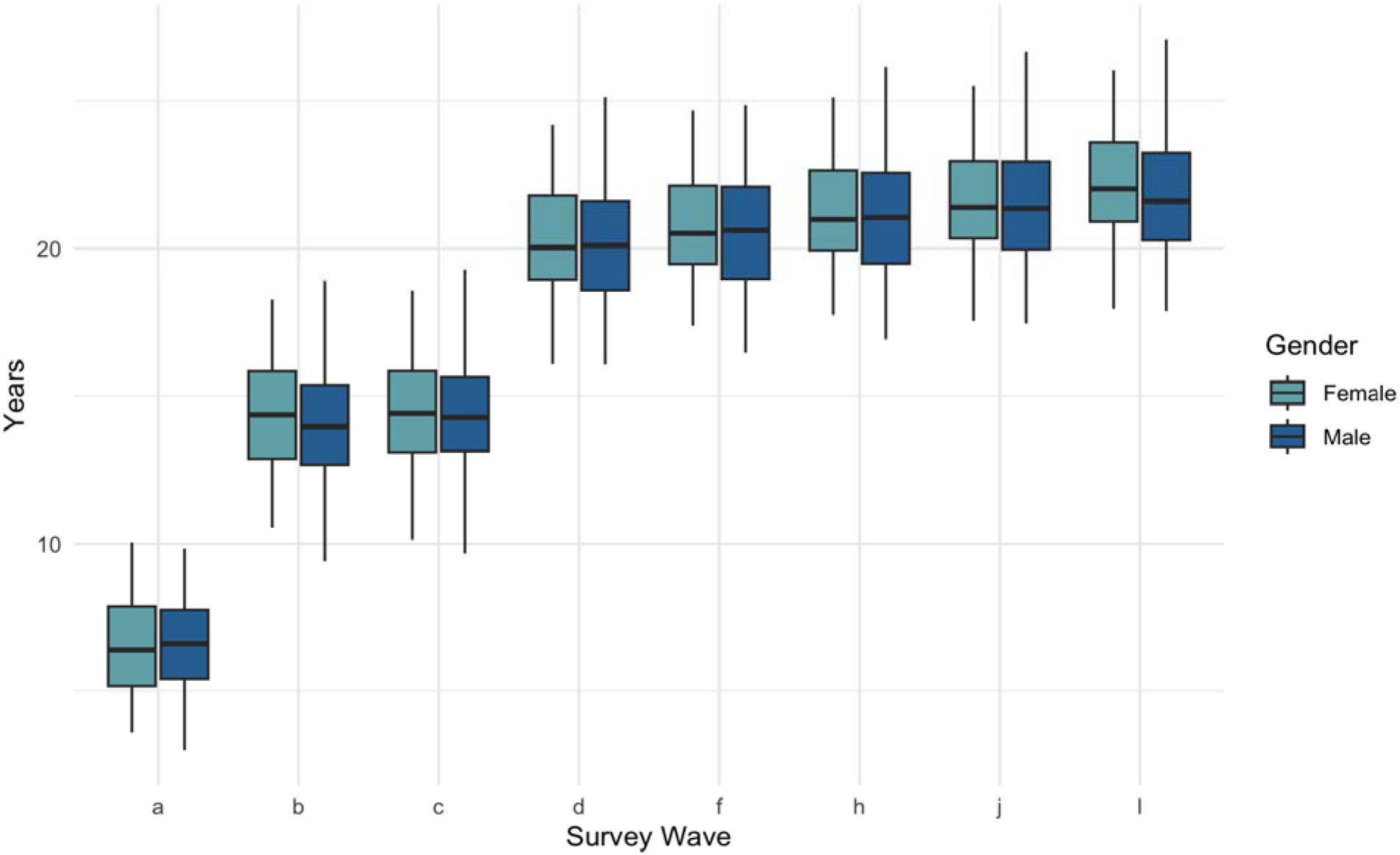
Distribution of participant ages by wave and sex.

**Figure S3.**
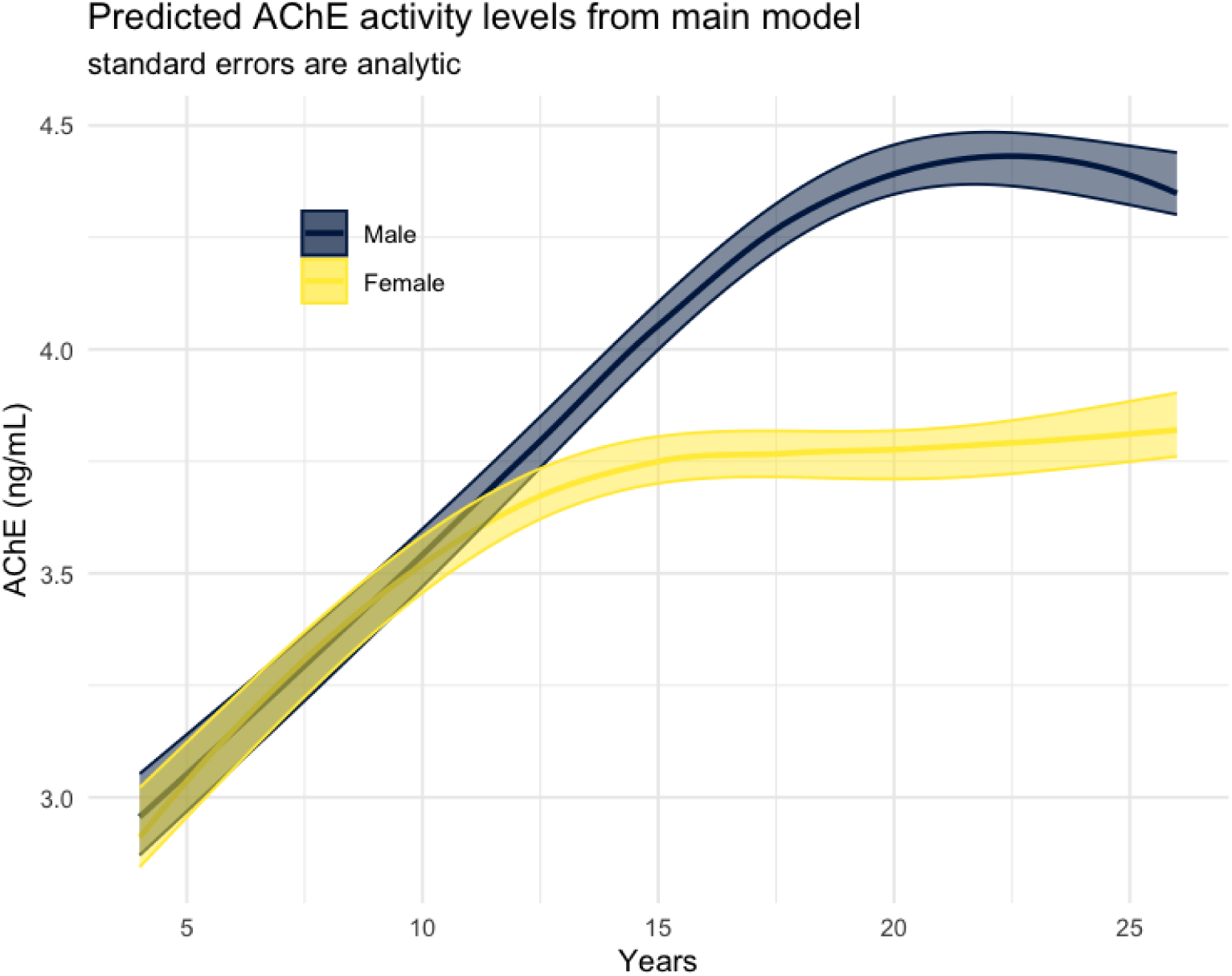
Predicted male and female AChE activity levels by age from main model (not bootstrapped confidence intervals).

**Figure S4.**
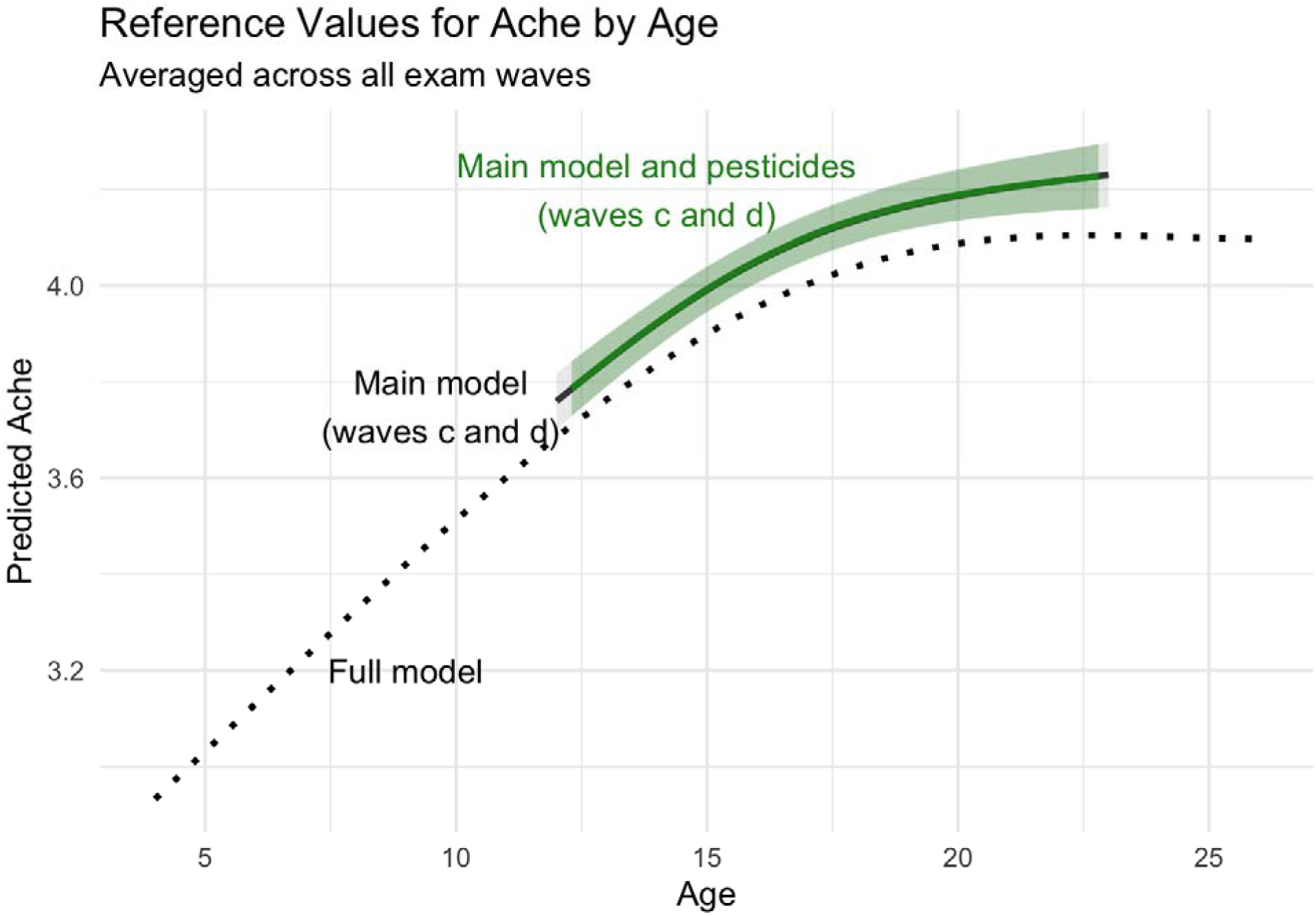
Results are robust to the inclusion of pesticide metabolites.

**Figure S5.**
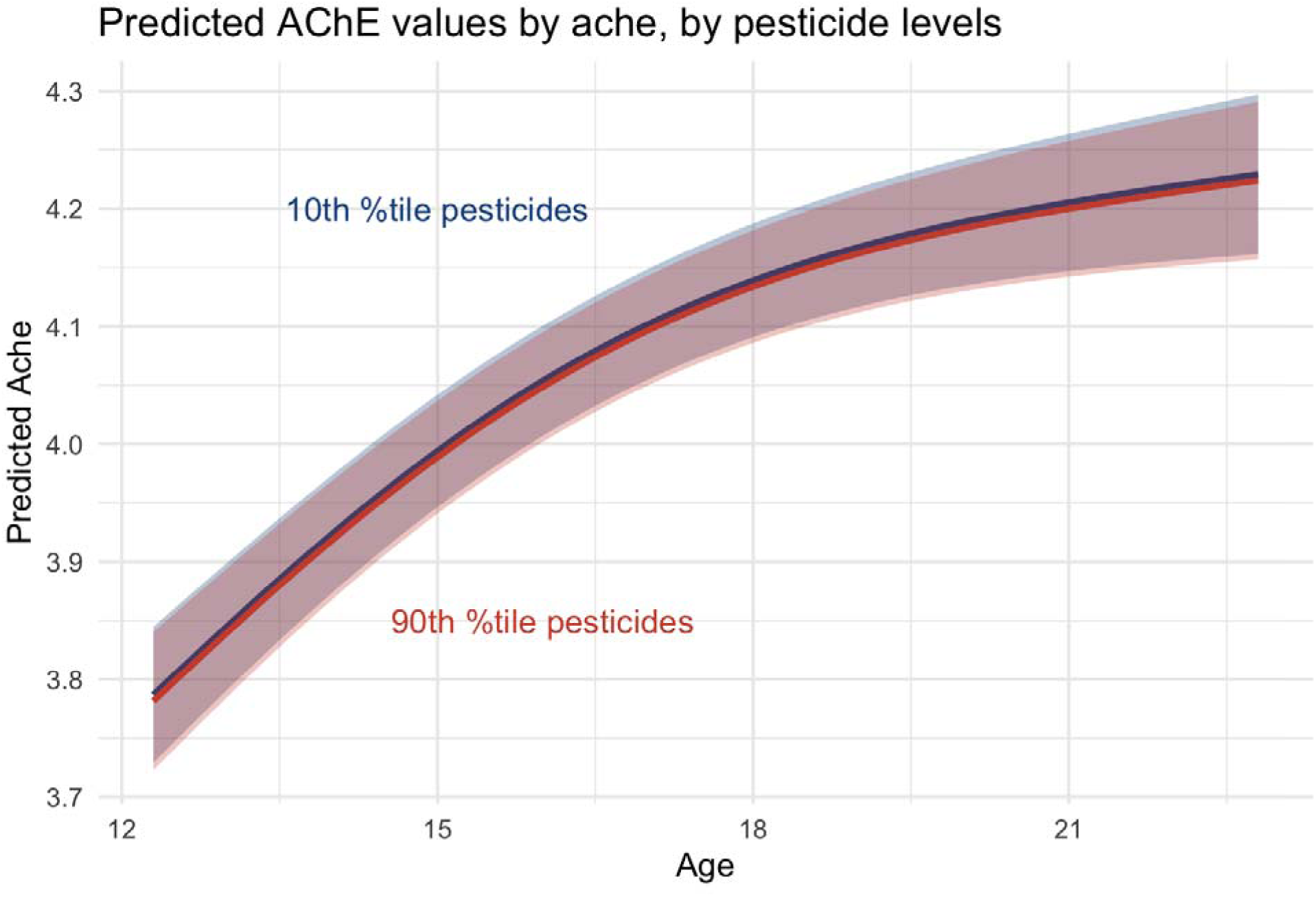
Predicted AChE activity levels are nearly identical when modeled at low and high pesticide levels.

## References

1. Taylor P. Anticholinesterase Agents. In: Brunton LL, Hilal-Dandan R, Knollmann BC, eds. Goodman & Gilman’s: The Pharmacological Basis of Therapeutics. 13th ed. McGraw-Hill Education; 2017. Accessed September 17, 2024. accessmedicine.mhmedical.com/content.aspx?aid=1162534099

2. Jaga K, Dharmani C. Sources of exposure to and public health implications of organophosphate pesticides. Rev Panam Salud Publica. 2003;14(3):171–185. doi:10.1590/S1020-49892003000800004

3. Colovic MB, Krstic DZ, Lazarevic-Pasti TD, Bondzic AM, Vasic VM. Acetylcholinesterase Inhibitors: Pharmacology and Toxicology. Current Neuropharmacology. 2013;11(3):315–335.

4. Arrieta DE, McCurdy SA, Henderson JD, Lefkowitz LJ, Reitstetter R, Wilson BW. Normal range of human red blood cell acetylcholinesterase activity. Drug and Chemical Toxicology. 2009;32(3):182–185. doi:10.1080/01480540902863440

5. Lefkowitz LJ, Kupina JM, Hirth NL, et al. Intraindividual Stability of Human Erythrocyte Cholinesterase Activity. Clinical Chemistry. 2007;53(7):1358–1363. doi:10.1373/clinchem.2006.085258

6. Maroni M, Colosio C, Ferioli A, Fait A. Biological Monitoring of Pesticide Exposure: a review. Introduction. Toxicology. 2000;143(1):1–118. doi:10.1016/S0300-483X(99)00152-3

7. Karlsen RL, Sterri S, Lyngaas S, Fonnum F. Reference values for erythrocyte acetylcholinesterase and plasma cholinesterase activities in children, implications for organophosphate intoxication. Scandinavian Journal of Clinical and Laboratory Investigation. 1981;41(3):301–302. doi:10.1080/00365518109092048

8. Suarez-Lopez JR, Jacobs DR, Himes JH, Alexander BH, Lazovich D, Gunnar M. Lower acetylcholinesterase activity among children living with flower plantation workers. Environmental Research. 2012;114:53–59. doi:10.1016/j.envres.2012.01.007

9. Worek F, Schilha M, Neumaier K, et al. On-site analysis of acetylcholinesterase and butyrylcholinesterase activity with the ChE check mobile test kit—Determination of reference values and their relevance for diagnosis of exposure to organophosphorus compounds. Toxicology Letters. 2016;249:22–28. doi:10.1016/j.toxlet.2016.03.007

10. Wilson BW, Arrieta DE, Henderson JD. Monitoring cholinesterases to detect pesticide exposure. Chemico-Biological Interactions. 2005;157-158:253–256. doi:10.1016/j.cbi.2005.10.043

11. Abdel Rasoul GM, Abou Salem ME, Mechael AA, Hendy OM, Rohlman DS, Ismail AA. Effects of occupational pesticide exposure on children applying pesticides. NeuroToxicology. 2008;29(5):833–838. doi:10.1016/j.neuro.2008.06.009

12. G. M. Higgins, J. F. Muaiz, L. A. McCauley. Monitoring Acetylcholinesterase Levels In Migrant Agricultural Workers and Their Children Using a Portable Test Kit. Journal of Agricultural Safety and Health. 2001;7(1):35–49. doi:10.13031/2013.2605

13. Crane AL, Abdel Rasoul G, Ismail AA, et al. Longitudinal assessment of chlorpyrifos exposure and effect biomarkers in adolescent Egyptian agricultural workers. J Expo Sci Environ Epidemiol. 2013;23(4):356–362. doi:10.1038/jes.2012.113

14. Galea KS, MacCalman L, Jones K, et al. Urinary biomarker concentrations of captan, chlormequat, chlorpyrifos and cypermethrin in UK adults and children living near agricultural land. J Expo Sci Environ Epidemiol. 2015;25(6):623–631. doi:10.1038/jes.2015.54

15. Gamlin J, Romo PD, Hesketh T. Exposure of young children working on Mexican tobacco plantations to organophosphorous and carbamic pesticides, indicated by cholinesterase depression. Child. 2007;33(3):246–248. doi:10.1111/j.1365-2214.2006.00702.x

16. Thompson B, Griffith WC, Barr DB, Coronado GD, Vigoren EM, Faustman EM. Variability in the take-home pathway: Farmworkers and non-farmworkers and their children. J Expo Sci Environ Epidemiol. 2014;24(5):522–531. doi:10.1038/jes.2014.12

17. Suarez-Lopez JR, Jacobs DR, Himes JH, Alexander BH, Lazovich D, Gunnar M. Lower acetylcholinesterase activity among children living with flower plantation workers. Environmental Research. 2012;114:53–59. doi:10.1016/j.envres.2012.01.007

18. Suarez-Lopez JR, Hong V, McDonald KN, Suarez-Torres J, López D, De La Cruz F. Home proximity to flower plantations and higher systolic blood pressure among children. International Journal of Hygiene and Environmental Health. 2018;221(8):1077–1084. doi:10.1016/j.ijheh.2018.08.006

19. Suarez-Lopez JR, Butcher CR, Gahagan S, Checkoway H, Alexander BH, Al-Delaimy WK. Acetylcholinesterase activity and time after a peak pesticide-use period among Ecuadorian children. Int Arch Occup Environ Health. 2018;91(2):175–184. doi:10.1007/s00420-017-1265-4

20. World Health Organization. Training Course on Child Growth Assessment. 2008;7(0):25–36.

21. World Health Organization Multicentre Growth Reference Study Group. World Health Organization Child Growth Standards based on length/height, weight and age. Acta paediatrica. 2006;450:76–85.

22. EQM Research Inc. Test-mate ChE Cholinesterase Test System (Model 400). Instruction Manual. 2003. http://www.eqmresearch.com/Manual-E.pdf

23. Arrieta DE, McCurdy S a, Henderson JD, Lefkowitz LJ, Reitstetter R, Wilson BW. Normal range of human red blood cell acetylcholinesterase activity. Drug and chemical toxicology. 2009;32(3):182–185. doi:10.1080/01480540902863440

24. Lefkowitz LJ, Kupina JM, Hirth NL, et al. Intraindividual stability of human erythrocyte cholinesterase activity. Clinical chemistry. 2007;53(7):1358–1363. doi:10.1373/clinchem.2006.085258

25. Magnotti RA, Dowling K, Eberly JP, McConnell RS. Field measurement of plasma and erythrocyte cholinesterases. Clinica Chimica Acta. 1988;176(3):315–332. doi:10.1016/0009-8981(88)90190-8

26. van der Hoek W, Konradsen F, Athukorala K, Wanigadewa T. Pesticide poisoning: A major health problem in Sri Lanka. Social Science & Medicine. 1998;46(4-5):495–504. doi:10.1016/S0277-9536(97)00193-7

27. Al-Shatti a K, El-Desouky M, Zaki R, Abu Al-Azem M, Al-Lagani M. Health care for pesticide applicators in a locust eradication campaign in Kuwait (1988-1989). Environmental research. 1997;73(1-2):219–226. doi:10.1006/enrs.1997.3735

28. Innes DF, Fuller BH, Berger GMB. Low serum cholinesterase levels in rural workers exposed to organophosphate pesticide sprays. South African Medical Journal. 1990;78(10):581–583.

29. Jeyaratnam J, Lun KC, Phoon WO. Blood cholinesterase levels among agricultural workers in four Asian countries. Toxicology letters. 1986;33(1-3):195–201.

